# Normalized barriers and unaddressed concerns: A qualitative study of the lived experiences of adults living in rural areas with advanced chronic kidney disease

**DOI:** 10.64898/2026.07.05.26356878

**Authors:** Gabriela Sanders, Ishaan Kumar, Anne Dade, Steven L. Bernstein, Clay A. Block, Hannah Crowe-Cumella, Glyn Elwyn, Lorraine Gerraughty, Vanessa Junkins, Annika Milliman, Joseph P. Nano, Niveditta Ramkumar, Ailyn Sierpe, Quenton Turner-Gee, Ann O’Hare, JoAnna K. Leyenaar, Catherine H. Saunders

**Affiliations:** Dartmouth Health; The Dartmouth Institute for Health Policy and Clinical Practice, Geisel School of Medicine at Dartmouth College; University of California, San Francisco; University of Washington

## Abstract

**Rationale & Objective:** People who live in rural areas with advanced chronic kidney disease (CKD) face well-documented structural barriers to receiving care, yet little is known about how they experience their illness or perceive interactions with their healthcare teams. We aimed to characterize the lived experiences and care perceptions of adults living in rural areas with advanced, pre-dialysis CKD.

**Study Design:** We conducted semi-structured qualitative interviews with patients and care partners.

**Setting & Participants:** We recruited patients with advanced CKD (stages 4–5, not on dialysis) and their care partners from a single hospital-based nephrology clinic in northern New England serving a predominantly rural population.

**Analytical Approach:** We analyzed interview transcripts using participatory Practical Thematic Analysis (PTA), an inductive, stakeholder-engaged approach to qualitative analysis.

**Results:** We interviewed 12 patients and 4 care partners. Four themes were identified: (1) logistical challenges of rural CKD care were pervasive but frequently normalized as an expected feature of rural life; (2) disease progression and future treatments were sources of uncertainty and concern, with expectations about dialysis often shaped by peer accounts rather than clinical discussion; (3) clinical conversations centered on laboratory results and medications, leaving emotional concerns and psychologic challenges unaddressed; and (4) physical symptoms and lifestyle changes were common but frequently attributed to comorbid conditions rather than to CKD.

**Limitations:** Recruitment from a single clinic with a small, racially homogeneous sample limits transferability. While in-person recruitment may have excluded patients with greater transportation barriers, those who attended represent a population navigating substantial access challenges to receive care.

**Conclusions:** Adults living in rural areas with advanced CKD experience logistical, emotional, and informational challenges broadly consistent with those reported in non-rural CKD populations. Patients normalized geographic barriers and did not consistently identify rurality as a source of disadvantage, even as structural barriers persisted. These findings support the development of structured communication approaches in nephrology care that invite discussion of disease trajectory, daily life impacts, and emotional concerns.

## Background

Chronic kidney disease (CKD) is a progressive condition that disproportionately affects rural communities, where people experience both higher disease burden^1,2^ and persistent barriers to care.^3^ In rural settings, limited access to specialists and laboratory services, long travel distances to appointments and pharmacies, and transportation-related costs contribute to financial strain and missed work.^4–8^ Prior studies have documented lower rates of guideline-concordant CKD care and higher CKD-related mortality among rural populations compared to those living in urban areas.^9,10^

For people with advanced CKD (stages 4–5 CKD, not requiring dialysis),^11^ care typically involves intensive monitoring, medication adjustments, and discussions related to disease progression and future treatment planning, which requires effective clinical communication.^12,13^ Much pre-dialysis CKD management occurs outside of clinical settings, such as diet modifications and blood pressure monitoring, making self-management an important topic of discussion during visits.^14,15,16^ Prior studies have described CKD encounters as clinician-led and marked by complex medical language, which may limit patient participation.^17,18^ Yet, whether and how these dynamics shape the experiences of patients living in rural areas with advanced CKD remains unknown.

Despite well-documented rural-urban disparities in CKD care and outcomes, little is known about how people with advanced, pre-dialysis CKD in rural U.S. settings experience their illness or perceive their care. Prior qualitative research has described people’s lived experiences with CKD as shaped by lifestyle disruption, financial burden, and fear of disease progression,^7, 14^ but much of this work has been conducted outside of the U.S. or without a specific rural context.^4–7, 10^ Additionally, qualitative research has more often focused on patients receiving dialysis rather than those in the pre-dialysis period,^7^ despite this being a time when engagement with healthcare systems intensifies and patient-centered communication becomes increasingly important.^12,19^

To address these gaps, we conducted a qualitative study to characterize the lived experiences and care perceptions of adults living in rural areas with advanced, pre-dialysis CKD.

## Materials and Methods

### Design

We conducted a qualitative study using semi-structured interviews, nested within a larger study (K01DK139400, PI Saunders) aimed at developing and testing a clinical agenda-setting tool for patients with advanced CKD in a rural-serving nephrology clinic. Findings related to agenda-setting will be reported separately. We adhered to the Consolidated criteria for Reporting Qualitative Research (COREQ) (Appendix).^20^ Our Community Advisory Board (CAB) consisted of three people living with kidney disease and two care partners, who were engaged on this study from design through dissemination, offering perspectives on study developments, findings, and interpretations. The Dartmouth Health Institutional Review Board approved this study in October 2024 (STUDY02002598).

### Participants

We recruited purposively, seeking variation in gender, socioeconomic status, employment, and educational background to capture a range of perspectives. We defined advanced CKD as stage 4 (estimated glomerular filtration rate [eGFR] 15–29 mL/min/1.73 m²) or stage 5 (eGFR <15 mL/min/1.73 m²).^11^

Eligible patients were 1) 18 years or older, 2) English-speaking, 3) able to provide informed consent, and 4) attending an outpatient visit at a hospital-based nephrology clinic in Northern New England, where our study took place. Patients were excluded if they were currently receiving dialysis. Care partners were also eligible to participate in a joint interview with patients.

While not an eligibility criterion, rural residence was assessed for all participants - including care partners - using Rural-Urban Commuting Area (RUCA) codes of 4 or higher, assigned to participant home addresses using a validated classification tool.^21,22^ Rural-Urban Commuting Area codes were developed by the United States Department of Agriculture Economic Research Service and categorize census tracts based on population density and commuting patterns.^23^

### Screening and recruitment

Patient eligibility was determined through electronic health record (EHR) chart review ahead of appointments. Of 157 patients identified as eligible through EHR chart review, 12 consented and completed interviews. Six patients were lost to follow up after unsuccessful contact attempts following their clinic visit. Enrollment was limited primarily by researcher availability for on-site recruitment rather than patient refusal or ineligibility. A researcher approached eligible patients — and care partners, if present — to explain the study, obtain informed consent, and schedule an interview. Interviews were conducted on the same day as clinic appointments, immediately before or after the encounter, or by video at a mutually agreed upon time.

### Procedures

Interviews were conducted by five researchers: AD, AS, GS, JN, and CHS. AD is a White female research associate with a master’s in public policy (MPP); her training in participatory qualitative methods and experience as an end-of-life doula informed her approach to patient-centered research. AS is a female health services researcher and PhD student with a master’s in social research methods (MSc); her work in participatory methods informed her analytic approach. GS is a White female MD candidate whose background in rural healthcare delivery and palliative care informed her interpretation of study data. JPN is a White male clinical research coordinator with an MPH and a master’s in quantitative biomedical sciences. CHS is a White female health services researcher with an MPH and PhD in health policy and clinical practice; her experience living with a chronic health condition informed her interpretation of participant accounts. Some team members preferred not to report race and ethnicity. The research team is predominantly White, consistent with the study sample, and this shared cultural context is acknowledged as a potential influence on both data collection and interpretation.

No prior relationship existed between the research team and study participants. Participants were informed that researchers were affiliated with Dartmouth Health and were conducting a study on the lived experiences of adults with advanced CKD.

We audio-recorded interviews with participant consent and had recordings professionally transcribed verbatim. Field notes were not systematically recorded during this study. Patient participants received $25 gift cards. Interview duration ranged from 45 to 90 minutes. There were no repeat interviews. For participants who provided email addresses, we offered a copy of the transcript; no participants requested the transcript.

### Data collection instruments

We developed and piloted a semi-structured interview guide with input from our CAB. The guide explored the needs and experiences of people with advanced CKD, with particular attention to rural-specific experiences including travel to appointments and medication access challenges.^24^ The guide also included questions about clinical visit agenda-setting, which are not the focus of this paper but are relevant to the larger study context, which will be reported separately. The interview guide for patients and care partners is available in Appendix 2.

Prior to interviews, patient participants completed a brief demographic survey (Appendix 3) and were screened for health literacy using the Single-Item Health Literacy Screener.^25^

### Analysis

#### Practical thematic analysis

We used an inductive approach to analyze interview transcripts using participatory Practical Thematic Analysis (PTA), a method selected to support iterative theme development grounded in participants’ accounts while formally incorporating stakeholder perspectives throughout analysis.^26–30^

Two trained coders (GS and IK) independently read all transcripts in full to support familiarization with the data and document initial impressions. IK is a male MD candidate with an MPH and a background in health services research. Both GS and IK acknowledge that their ongoing medical training shaped their initial orientations to the data. Prior to coding, both coders engaged in reflexive discussion with the senior qualitative investigator (CHS) to surface assumptions and potential interpretive biases.

GS and IK then independently coded a subset of transcripts using <u>ATLAS.ti</u> and met iteratively to develop a negotiated shared codebook. The codebook was refined continuously throughout early coding to incorporate emergent and recurring codes. Once stabilized, both coders independently applied the shared framework to the remaining transcripts. Thematic saturation was assessed throughout data collection and analysis, with attention to both repetition across transcripts and sufficient depth of coverage within themes. No new themes emerged after the twelfth interview. The codebook is available from the corresponding author upon reasonable request.

Following coding, GS and IK independently reviewed the full dataset, and each developed a draft thematic model summarizing key patterns across codes. Both coders then presented their draft models at a PTA-style thematic analysis session facilitated by CHS. Two CAB members - one patient (QTG) and one care partner (NR) - participated as collaborative partners. The group identified areas of commonality and disagreement, and constructed a shared model through facilitated discussion.^30^ CAB involvement at this stage supported the validity and contextual grounding of the findings.^30^

Following the thematic analysis session, the analysis team (GS, IK, CHS) refined theme definitions, verified that each theme was well supported across transcripts, and attended to disconfirming evidence. A final thematic model was developed with representative quotations labeled by participant identifiers.

## Results

### Characteristics of the sample

#### Demographics and metrics

We interviewed 12 patients and 4 care partners. Demographic data were collected from patient participants only, as the study’s focus was on patient experiences and care perceptions.

Among the 12 patient participants, eight were male and four were female (Table 1). The majority were 55 years or older (n=10, 83.3%) and had attained a high school diploma or less (n=9, 75%). All patient participants identified as White or Caucasian (n=12, 100%), consistent with the majority White population in Northern New England.^31,32^ All patient participants (n=12, 100%) resided in rural areas, as defined by Rural-Urban Commuting Area (RUCA) codes of 4 or higher.^22^

**Table 1.** Demographic characteristics of patient participants.

|  | Number of patient participants<br>N=12 |
| --- | --- |
| **Age** |  |
| 35-54 years old | 2 |
| 55-74 years old | 6 |
| 75+ | 4 |
| **Gender** |  |
| Female | 4 |
| Male | 8 |
| **RUCA code** |  |
| 4-6 (Large rural) | 5 |
| 7-9 (Small rural) | 2 |
| 10 (Isolated rural) | 5 |
| **Healthcare access difficulty<sup>†</sup>** |  |
| No | 10 |
| Yes | 0 |
| **Education** |  |
| Less than high school | 3 |
| High school diploma or equivalent | 6 |
| Some college, Associate's or Bachelor's degree | 3 |
| **Insurance<sup>†</sup>** |  |
| Medicaid | 3 |
| Medicare | 4 |
| Private insurance | 4 |
| **Comfort with medical forms (health literacy)<sup>†</sup>** |  |
| No comfort/ little comfort | 4 |
| Quite a bit/ extremely comfortable | 7 |
| **Employment** |  |
| Full-time | 3 |
| Part-time | 2 |
| Unemployed | 2 |
| Retired | 5 |

† Missing data: healthcare access difficulty (n=2), insurance (n=1), health literacy (n=1)

### Theme 1: Logistical challenges of rural CKD care were pervasive and often normalized

*Subthemes: Reliance on care partners for transportation and access; Telehealth as a means of maintaining continuous care; and Pharmacy-related barriers to medication access*

Although many patients lived substantial distances from the nephrology clinic, most described travel as a normal and accepted part of rural life. Long drives, scheduled time away from work, and distance to pharmacies were not consistently framed as barriers to care, even when they created real disruptions. Patient 33 summarized, “*One of the things that you should consider is when you live rurally, you plan that it takes time to go places. So it doesn’t necessarily impact everything because that’s the norm.*”

Patients identified several strategies that helped manage the logistical burden of rural CKD care, including telehealth, mail-order pharmacies, and care partner support.

#### Reliance on care partners for transportation and access

Transportation was one of the clearest ways rurality shaped CKD care. Many participants did not feel comfortable driving long distances themselves and relied heavily on family and close friends for rides to appointments. For some patients, this reliance sometimes led to missed appointments and difficulties getting consistent CKD care. As Patient 143 explained, “I *had to reschedule my last nephrology [appointment] because I couldn’t get a ride*.”

For patients without these social networks, the burden was even greater. Patient 16 described, *“It’s starting to get hard [to get] down here…driving by yourself two hours, two and a half hours, one way.”*

These transportation challenges reflected a broader pattern of daily management supported by care partners. Beyond rides, many care partners took responsibility for medication refills, appointment schedules, and paperwork. As Patient 30 described, “*My wife takes care of all my medications. Without her, I’d be lost.”*

For some care partners, their responsibility for transportation and supporting patients’self-management created an additional and sometimes unacknowledged workload.

Patient 111’s family member shared, *“It puts a lot of extra work on me, you know extra responsibilities, I should say. So it does get kind of hard to try and juggle everything and do, you know, everything.”*

#### Telehealth as a means of maintaining continuous care

Telehealth emerged as an important modality for maintaining continuous CKD care when weather or transportation prevented patients living in rural areas from attending in-person visits. Patient 151 shared, “*[I have missed appointments due to snow]. But I’m fortunate with telehealth to be able to switch things over.”*

For patients dependent on care partners for rides, telehealth allowed for regular contact with their clinicians. As Patient 143 explained, *“If I don’t have a ride, my [care partner] would rather me do a telehealth visit because a lot of times he can’t, you know, get me there because he has his own health problems.”*

#### Pharmacy-related barriers to medication access

Medication access was shaped by a combination of travel distance, limited local pharmacy capacity, and insurance-related requirements, with mail-order prescriptions serving as both a practical solution and an additional logistical consideration. Several patients described disruptions related to medication availability and pharmacy logistics.

Patient 151 outlined how the structural barriers of rural care affected his ability to access his CKD medications: *“[The first pharmacy] is a half hour from where I live. Some medications I can’t get locally… then I use [the second pharmacy], and I went six weeks without medication just recently, but there’s been issues with that pharmacy… that I was told because they’re a small town pharmacy…It’s very disruptive. But I found a way around it. I need to go to [the third pharmacy] which is another 20 minutes down the road.”*

Mail-order delivery was also described as part of this medication access landscape. Use of mail-order prescriptions reflected a mix of insurance requirements, cost considerations, and travel burden. As Patient 110 explained, *“Our insurance offers a mail order, and the 90 days basically costs almost the same as a monthly prescription…we get our meds at the end of our driveway. So we can check it every single day, twice a day if a…strong medication is coming’”*

### Theme 2: Disease progression and future treatments were sources of uncertainty and concern

*Subthemes: Dialysis as the “end of the road” and fears of dying; and Dialysis disrupting daily life*

Patients described a sense of uncertainty and fear about how their CKD would progress. For those with few physical symptoms, the absence of bodily feedback made disease progression feel abstract and difficult to anticipate. Expectations about dialysis, the most commonly discussed future treatment, were frequently shaped by conversations with peers rather than by clinical discussions, and carried significant emotional weight.

#### Dialysis as the “end of the road” and fears of dying

For many participants, dialysis symbolized a major decline in health and was a marker for a loss of independence and an increased chance of dying. Many beliefs and expectations of dialysis were shaped by secondhand accounts rather than clinical discussions. Patient 16 simply stated, “*I had a friend that had dialysis. He didn’t last long.”*

Limited clinical understanding further contributed to this uncertainty about dialysis and disease progression. Several participants described a vague sense of what dialysis entails and filled in the gaps with stories from others. Patient 143 explained, “*I really don’t have knowledge of dialysis…you know you take the blood out and wash it and all that. I don’t really know the details. People tell me that’s the end of the road.”*

Fear and uncertainty around disease progression and life expectancy were common, even when patients could not clearly articulate what progression would look like for them. When prompted about their biggest worry, Patient 8 shared, “*Oh, that I’m not going to live for another 10 years.”* Some participants described understanding advanced CKD progression as involving a narrow set of possible outcomes. When sharing fears about his kidney disease progression, Patient 16 stated, *“You know what happens after the end of stage five? Two things can happen: death or go on to dialysis.”*

Transplantation was occasionally raised by participants, though it was typically perceived as uncertain or distant rather than an actively sought alternative. Patient 33 questioned his own eligibility, framing transplant alongside dying as a possible but uncertain outcome, sharing his fear of *“having to get a transplant, or even if I’m able to get a transplant.“*

#### Dialysis disrupting daily life

Patients feared how dialysis might reshape their daily routines and sense of identity. Concerns focused on the time commitment, physical toll, finances, and potential loss of employment or means of enjoyment.

Patient 30 anticipated the prospect of losing his job, stating, “*I have to give up my line of work… It’s automatic federal law that once I go on dialysis, I can’t drive commercial vehicles anymore.”* Other participants commented on the travel distance and frequency of appointments disrupting their hobbies, relationships, and finances. Speaking to the financial strain, Patient 16 shared, *“I’ll have to do dialysis and then that’s three times a week. Well, that’s going to be a pain in the back. That’s going to be costly. I’m a poor man to start with.”*

### Theme 3: Clinical conversations often centered on lab results and medications, leaving emotional concerns unaddressed

*Subthemes: Trust in clinician-directed care; Preference for straightforward communication; Unspoken emotional concerns; and Centering patient values and self-advocacy*

Participants described strong trust in their nephrology teams and valued direct, clear communication. Clinical visits were experienced as structured around laboratory results and medication management, which participants generally accepted. However, this structure left limited space for emotional concerns, which patients often chose not to raise.

#### Trust in clinician-directed care

Patients frequently deferred to their clinicians when making treatment decisions, describing confidence in their team’s training and judgement, which was underscored by their long-standing relationships. For most participants, trust translated into willingness to follow clinical guidance, though this did not reflect a preference for being excluded from decisions. As the spouse of Patient 31 described, “*I think that’s why my husband’s doing so well, because he follows them to the letter. If they say, “Oh, you can’t have this anymore,” he won’t have it.”*

Similarly, Patient 30 explained, “*I’m comfortable with them. They’re the ones that train for this. I’m living with it, but I trust them.”*

#### Preference for straightforward communication

Participants consistently valued direct clinical communication, especially when discussing difficult information. They viewed straightforwardness as a sign of honesty and transparency, creating a shared understanding between themselves and their clinicians. Patient 26 shared, “*I like people that are straightforward and honest, whether it’s brutal or not…I’d rather have honesty. That’s important to me.”*

Others expressed the frustrations of overly long and vague explanations. As Patient 8 said, “*If I have a sickness, I want to be told I have a sickness, and I want to be told the facts. I want to be upfront. You know I don’t want to sit back and listen to somebody talk to me for an hour and really not tell me anything.”*

#### Unspoken emotional concerns

Although many patients felt comfortable with their clinicians, some described hesitancy in raising emotional or psychological concerns. Two reasons emerged: not wanting to appear fragile, and a belief that emotional topics fell outside the appropriate scope of nephrology care. As Patient 29 explained, *‘I got aches and pains like any other old man, but what are you going to do? [I am not talking about them in a visit] because they’re all extracurricular activities,”* suggesting that sharing his feelings of discomfort is outside of the scope of a clinical visit.

Others described avoiding topics that felt too personal or emotional. Patient 143 reflected, *“I don’t enjoy life at all…I don’t usually talk about it. I mean, if [the clinical team asks] how’s my day going? I’ll say, you know, honestly, how I’m doing. But, I mean, we never really talked about how life is, you know, crappy at times…I feel like it’s more emotional than, you know, anything else.”*

#### Centering patient values and self-advocacy

While many patients deferred to clinician-guided medical direction, some described instances where their own preferences guided medical decisions. A care partner for Patient 31 recounted a decision point around dialysis, sharing, “*[My husband’s physicians] said [my husband] should be on dialysis, but he chose not to. That should be the patient’s choice, which the doctors totally respect.”*

Patients also reflected on learning to advocate for themselves over time. As Patient 151 shared, “*When I don’t understand things, I tend to go like, ‘Okay. What’s that?’ And that’s something I had to learn…But five years into it, I know more about to ask and not to smile and nod and walk away.”*

### Theme 4: Physical symptoms and lifestyle changes were common but often attributed to comorbid conditions rather than CKD

*Subthemes: Fatigue and its impact on daily functioning; Dietary restrictions as an ongoing reminder of CKD; and Attribution of symptoms to comorbid conditions*

Many participants initially described CKD as a condition that existed in the background of their daily lives. As Patient 6 reflected, *“I don’t feel like I have kidney disease. I really don’t. I mean, I know I do, and I know I have to watch things, but I do what I want when I want.”*

With continued discussion, however, participants described substantial physical and lifestyle burdens associated with CKD. Fatigue and dietary restrictions shaped their daily routines, social activities, and relationships, even if participants did not immediately link these changes to their CKD. Some participants attributed physical symptoms to other comorbid conditions, which reinforced the sense that CKD was not always felt directly in the body.

#### Fatigue and its impact on daily functioning

Participants frequently described fatigue as one of the most disruptive aspects of their health, even when they did not directly connect it to CKD. Fatigue limited their ability to work, maintain social relationships, or engage in meaningful activities.

Others spoke about how fatigue affected their ability to enjoy activities and passions, particularly outdoor activities associated with rural living. Patient 111 and his care partner shared, *“You used to grow a garden and we had a big garden. It has changed our life. As [CKD has] advanced, we’ve had to give up a lot of our normal activity. I used to do a lot of stuff. Now, I don’t. I can hardly do anything.”*

Patient 8 similarly described, *“I’m not as active as I used to be. I can’t go out and cut trees down. And you know I can’t raise cattle anymore. I had to get rid of them.”*

#### Dietary restrictions as an ongoing reminder of CKD

Dietary restrictions were one of the most concrete reminders of CKD in daily life, often requiring lifestyle adjustment. Patient 6 described reading every food label, noting that tracking his sodium intake was a reminder of CKD. Others explained how diet changes gradually became more routine through the course of their disease. Patient 31 shared, *“You had to watch the sodium, watch the potassium. But now it’s gotten to be like second nature. I used to be a big salt eater. No salt anymore. It took me a while to get used to it, I’ll tell you.”*

#### Attribution of symptoms to comorbid conditions

Some participants attributed the physical symptoms of CKD to other chronic conditions rather than to CKD, which contributed to a sense that CKD itself was not impacting their daily experiences. Others recognized the interconnectedness of their illnesses.

As Patient 151 explained, *“I’ve got comorbidities like type 2 diabetes to go with it…So I don’t specifically look at it as like this has to do with chronic kidney. I am a ball of comorbidities together. And… if my sugar’s bad, that affects my kidneys.*”

Other participants were unsure which symptoms could be directly attributed to their different conditions. Patient 29 and his care partner debated the source of symptoms. Their care partner noted, *“He goes to the bathroom a lot… He does have mood swings, but I guess that’s with the sugar.”* Patient 29 then added, *“Yeah. Well, maybe it’s kidney. I don’t know.”*

## Discussion

### Summary of results

This study illustrates that adults living in rural areas with advanced CKD are largely accepting of the logistical challenges of accessing care, draw on the experiences of peers when forming expectations about disease progression, notice that clinical encounters focus on tests and medications more than psychosocial concerns, and tend to attribute symptoms to comorbid conditions rather than to CKD. Across these themes, care partners emerged as central figures, absorbing logistical burdens, filling informational gaps, and supporting daily disease management. Experiences varied meaningfully based on the availability of social support, with those lacking strong networks facing greater challenges across all domains.

### Results in context

Prior research has documented rural-urban disparities in CKD care and outcomes, though much of this work has focused on structural and epidemiologic differences rather than patients’ lived experiences.^1–10^ Despite probing on rural-specific concerns, the themes that emerged in our study were broadly consistent with qualitative descriptions of advanced CKD in non-rural settings, suggesting that the core challenges of living with advanced CKD are not uniquely rural in origin. Participants did not consistently describe rurality as a barrier but as an expected feature of everyday life. Long travel distances, time away from work, and limited pharmacy access were framed as inevitable and expected features of living rurally. However, the degree to which these logistical challenges posed real difficulties depended on the availability of care partners and social support, extending prior research by demonstrating that geographic barriers are not experienced uniformly.^4–8^ Consistent with prior research identifying telehealth as a strategy to extend nephrology care to geographically isolated populations,^33^ participants described telehealth as an important means of maintaining continuous care when weather or transportation prevented in-person attendance.

Uncertainty about disease progression and future treatments was a central concern across interviews. Prior qualitative literature has shown that patients with advanced CKD often feel unprepared for dialysis,^4–7,12,19^ and our findings extend this by highlighting the emotional and symbolic meaning attached to it. Dialysis was frequently understood not only as a treatment option but as a marker of decline, loss of independence, and proximity to death.

Transplantation, when mentioned, was framed as an improbable outcome, suggesting that patients have limited awareness of their own eligibility. Expectations around progression were often shaped by secondhand accounts rather than clinical discussions, suggesting that patients fill informational gaps through social networks.

Clinical communication in CKD has been described as clinician-led and heavily focused on laboratory values and medication management.^17,18^ Our findings are consistent with this characterization. Participants valued direct, straightforward communication and expressed strong trust in their nephrology teams, often deferring to clinical expertise. However, similar to prior work,^34^ emotional and existential concerns were not consistently integrated into routine clinical conversations. Patients’ hesitancy to raise these concerns was related to beliefs about what falls within the scope of nephrology care, suggesting that even in trusting clinical relationships, the structure of visits may inadvertently signal that emotional concerns are not welcome.

Participants described CKD as existing in the background of daily life, even as fatigue, dietary restrictions, and reduced physical capacity disrupted their routines. The mismatch between CKD symptoms and patients’ ability to attribute them to the disease is consistent with prior descriptions of CKD as an “invisible” condition.^14,35^ Our findings reinforce this characterization, showing that the presence of comorbid conditions, particularly diabetes and hypertension, provided alternative explanations for symptoms that further obscured the role of CKD in patients’ daily experiences.^35,36^

### Implications

These findings have implications for clinical practice in nephrology, particularly in rural-serving settings. Although participants normalized geographic barriers, logistical challenges - particularly transportation dependence and limited social support - remained consequential.

Health systems should consider practical solutions including transportation benefit programs and expanded telehealth infrastructure to reduce access disparities for patients with limited social networks.

Structured communication approaches during clinical encounters represent a particularly important opportunity. Clinicians could proactively introduce discussions about disease trajectory, dialysis transition, and life expectancy and revisit them over time. Structured agenda-setting tools that explicitly invite discussion of daily life impacts may help prevent informational gaps from being filled through secondhand accounts and signal that these concerns are within the appropriate scope of nephrology care.^37^ Integrating serious illness communication and palliative care training into nephrology practice may further support these conversations.^38, 39^

Because participants frequently attribute CKD symptoms to comorbid conditions, clinicians may need to more explicitly connect physical changes to CKD itself. This may support patients in recognizing CKD as an active contributor to their daily experiences and encourage more complete disclosure during visits.

Given that participants relied heavily on peer accounts to understand disease progression and future treatments, peer support programs connecting patients with others at similar disease stages may offer a more balanced perspective and create space for emotional concerns that patients withhold from clinical encounters.^40^

### Strengths and limitations

This study has several methodological strengths. Community Advisory Board members with lived experience of CKD contributed to study design, analysis, and interpretation, grounding the findings in participant perspectives. Iterative codebook development and independent coding by two trained analysts, followed by a collaborative PTA session incorporating CAB input, support the rigor and credibility of the thematic model.

Several limitations should be noted. Recruitment from a single rural-serving clinic in Northern New England with a small, racially homogeneous sample limits transferability, though this sample is consistent with the local demographics of the region.^31,32^ In-person recruitment may have excluded patients with greater transportation barriers or higher telehealth reliance; that said, patients who attended in-person appointments at a clinic requiring substantial travel themselves represent a population already navigating significant access and transportation challenges to receive care. Conducting interviews on the same day as clinical encounters and in proximity to the clinical environment may have introduced social desirability bias, particularly around participants’ perceptions of their care teams and willingness to disclose emotional concerns. Joint interviews with patients and care partners, while providing valuable dyadic perspectives, may have limited individual disclosure on sensitive topics including fear of dying, financial strain, and relationship burden. Interview duration of 45–90 minutes may have favored participation by those with more time, energy, or engagement, introducing additional selection bias.

## Conclusion

Adults living in rural areas with advanced, pre-dialysis CKD normalized the logistical challenges of rural living even as those challenges remained consequential, particularly for those with limited social support. The core difficulties participants described, including disease uncertainty, unaddressed emotional concerns, and clinician-led communication, were broadly consistent with qualitative accounts of advanced CKD in non-rural settings, suggesting these challenges are not uniquely geographic in origin. These findings support structured, patient-centered communication approaches in nephrology that invite discussion of disease trajectory, daily life impacts, and emotional concerns, regardless of geographic context.

## Data Availability

All data produced in the present study are available upon reasonable request to the authors

---

**Table 2:** Supplementary Quotes Table.

| Theme | Subtheme | Quotes |
| --- | --- | --- |
| Theme 1: Logistical challenges of rural CKD care were pervasive and often normalized. | Reliance on care partners for transportation and access | Patient 16: It's starting to get hard to get down here...Driving by yourself two hours, two and a half hours, one way. |
|  |  | Patient 143: I had to reschedule my last nephrology because I couldn't get a ride. |
|  |  | Patient 110: It's been pretty good, pretty easy with my wife's help. I don't drive very often. But we have other family members that want to happen to our vehicle or myself. Someone would help it, for sure. |
|  |  | Patient 26: Usually, my niece or my nephew. Sometimes one of my girlfriends does. My mother has taken me before. I can't drive on the interstate. |
|  |  | Patient 31: She drives me. I drive a little bit, but I don't drive much like I used to. |
|  |  | Patient 111: Well, I can't do a lot of stuff. He can't do a lot of stuff. For me, it's put a lot of extra work on me, you know extra responsibilities, I should say. So it does get kind of hard to try and juggle everything and do you know everything. As a family member. |
|  |  | Patient 31: My husband, he doesn't want anything to do with the computers. It's not his era. You know, so that's why I take care of all that stuff instead of I mean, like he said, if he had to do that himself, it wouldn't happen. |
|  |  | Patient 31: She does it all. Medication, his appointments. He gets very confused on things, so I find that's what I'm here for. |
|  |  | Patient 8: I fill out everything they sent. And I do print. I do look at everything. He doesn't. It's his. I know all his passwords and everything because I have to go on and do it all. |
|  |  | Patient 30: My wife takes care of all my medications. Without her, I'd be lost. |
|  | Telehealth as a means of maintaining continuous care | Patient 143: If I don't have a ride, my other half would rather me do a telehealth visit because a lot of times he can't you know get me there because he has his own health problems of his own. |
|  |  | Patient 143: It's rural. But I mean, yeah, it is a challenge, depending on the weather, you know or access to a car. But luckily, we have telehealth now. |
|  |  | Patient 151: It's weather-dependent. Yeah, in the winter, it's a huge difference...I'm fortunate with telehealth to be able to switch things over. |
|  | Pharmacy-related barriers to medication access | Patient 151: [The first pharmacy] is a half hour from where I live. Some medications I can't get locally... then I use [the second pharmacy], and I went six weeks without medication just recently, but there's been issues with that pharmacy... that I was told because they're a small town pharmacy...It's very disruptive. But I found a way around it. I need to go to [the third pharmacy] which is another 20 minutes down the road. |
|  |  | Patient 110: Our insurance offers a mail order, and the 90 days basically costs almost the same as a monthly prescription. So you're kind of crazy not to do it that way. And we get our meal at the end of our driveway. So we can check it every single day, twice a day if we need to if something you know strong medication is coming or something. So that's what we opt to do. |
|  |  | Patient 16: If I don't do mail order, it's 20 minutes away. One way. But you've been doing the mail order type of thing. That's what I'm trying to do this year because it's cheaper. |
| Theme 2: Disease progression and future treatments were sources of uncertainty and concern. | Dialysis as the "end of the road" and fears of dying | Patient 16: I worry. I told them that today. I just worry about how my bloodwork is this time. Is it worse?...So you know what happens after the end of stage five? Two things can happen. Death or go on to dialysis. |
|  |  | Patient 29: She's scared the shit out of me. Six, eight months ago. She sent me an appointment with somebody to talk about dialysis. And I'm like, "Say what?" |
|  |  | Patient 29: I don't know what I'm going to do with the information when they tell me you got to go on dialysis. I don't have that plan yet. My brain tells me it ain't going to happen. |
|  |  | Patient 143: And I don't want dialysis. I worry about that...I just feel like that's the last thing there is. But the nurse yesterday told me that it's not. I feel like that's like stage five, the end of the road usually. That's what I think. I guess I really don't have knowledge of dialysis. I know it just you know you take the blood out and wash it and all that. I don't really know the details. People tell me that's the end of the road. |
|  |  | Patient 8: Oh, that I'm not going to live for another 10 years. |
|  |  | Patient 26: Because you know they're in the back of my head. I truly do not want to go through dialysis. If I can skip that, totally. You know, the longer I could go without having a transplant, that's even better. |
|  |  | Patient 33: Well, yeah, them dying, you know having to get a transplant or even if I'm able to get a transplant. |
|  | Dialysis disrupting daily life | Patient 30: The only thing that is a drawback to dialysis, I have to give up my line of work when I don't work...It's automatic federal law that once I go on dialysis, I can't drive commercial vehicles anymore. |
|  |  | Patient 16: And then when I get on the kit, I'll have to do dialysis and then that's three times a week. Well, that's going to be a pain in the back. |
|  |  | Patient 16: I had a friend that had dialysis. He didn't last long. He seemed to have been tired more too. But he didn't last long. He died, probably like four or five years....I just feel like it's going to ruin my day...Because I like to get up in the morning and get something done. When it hits summertime, I'm not in my house. And I'd hate to sit around in my house. I'll be outside. I'll ride my motorcycle. I'm always doing yard work. |
|  |  | Patient 30: If there's a way to keep going without it, dialysis doesn't interest me at all. I don't think it interests a lot of people. Well, my mother went through it for five and a half years, three days a week, four hours every trip. You know if I could get out of that, I would. |
|  |  | Patient 30: I have limited financial resources, and I don't worry about it. Happy, go lucky. I mean, I can't produce what I haven't got. And if I'm turned down because I don't have any money, I'll live with it. |
| Theme 3: Clinical conversations often centered on lab results and medications, leaving emotional concerns unaddressed. | Trust in clinician-directed care | Patient 30: I'm comfortable with them. They're the ones that train for this. I'm living with it, but I trust them. |
|  |  | Patient 29: They tell me what I need to know. I do it. I don't know why. If I had a broken leg and it hurt, I would understand how to deal with it. It's not that I don't believe I don't have chronic kidney disease. I don't feel it. So I can't grasp it. |
|  |  | Patient 110: I am open for suggestions and who knows better than the doctors? |
|  |  | Patient 143: And then the previous physician just you know he tells me what I should do, and you know you know I follow what I can...I don't think I've ever disagreed with this physician because he was pretty knowledgeable and understanding and listens and just tells me what he suggests. You don't have to do what they say, but I just always know to go by what they say. |
|  |  | Patient 26: She's proactive and she has been right along and she's straightforward and I just love it because she said it's better to be ahead of the game and be ready for it than have something happen and you know you're not prepared. |
|  |  | Patient 8: I really don't know that much about it yet. But I mean, I have full confidence in what [the physician] is recommending. He didn't give us a detailed plan or anything...But he involved us in a lot of things. |
|  |  | Patient 31: And I think that's why my husband's doing so well, because he follows them to the letter. If they say, "Oh, you can't have this anymore," he won't have it. |
|  |  | Patient 33: You have to take the meds that they prescribe, and you have to trust in what they're doing. |
|  |  | Patient 26: And I tell them all the time I am blessed that I don't know why, but I feel like my doctors are awesome because my primary, my kidney doctor, even my dermatologist doctor, they all connect. You want to make sure I feel like I am actually a patient not in and out. |
|  |  | Care Partner of Patient 6: I mean, we've had such a great experience with nephrology. She's had a tremendous experience, and that's why some of the earlier questions that were asked, she's been able to lead a very normal life. She's been able to do everything that she wants. |
|  | Preference for straightforward communication | Patient 30: Well, if I'm doing something wrong, she flat out tells me I'm wrong. |
|  |  | Patient 30: Like I was telling you about being straightforward is the way I like it to the point. If I'm going to die next week, tell me. |
|  |  | Patient 8: I just want you to know I'm a basic simple guy. I don't need a lot of whipped cream...If I have a sickness, I want to be told I have a sickness, and I want to be told the facts. I want to be upfront. You know I don't want to sit back and listen to somebody talk to me for an hour and really not tell me anything. |
|  |  | Patient 26: I like people that are straightforward and honest, whether it's brutal or not...I'd rather have honesty that's important to me. |
|  | Unspoken emotional concerns | Patient 143: But, you know, and just being older, you know, in your 40s, you got to have aches and pains by now. But it's a good thing to talk about. Enjoying life, that's a definite thing because I don't feel like I do, actually, to be honest. I don't enjoy life at all, so...I don't usually talk about it. I mean, if they talk about, like, how's my day going? I'll say, you know, honestly, how I'm doing. But, I mean, we never really talked about how life is, you know, crappy at times. You know, it's more I feel like it's more emotional than, you know, anything else. |
|  |  | Care Partner of Patient 31: But with my husband, when he was first diagnosed with kidney disease, you could not talk to him about treatment and stuff because he got very emotional. It was a very, very emotional subject with him and the doctors...He doesn't want to read about it. I think it puts too much stress, anxiety on him if he starts reading a lot of stuff about kidneys. He knows he's got the disease. He's dealing with it. |
|  |  | Patient 151: As somebody who suffers from depression and you know I could be introverted, something like this without personal interaction is I wouldn't say dangerous, but if I feel shut down, I don't want to communicate. If you give me a piece of paper, I'm checking no. I'm saying I'm fine. I'm smiling and nodding. I've been giving every inclination that I'm okay because I've learned that as a [40 to 50]-year-old guy. I can handle myself and be quiet, be polite, and then go a long way. |
|  |  | Patient 26: This is why I didn't tell them until 2022 that I was at stage four because I didn't want that...I didn't want [my family] to treat me like I'm frail. You know what I mean? I wanted to still be treated like they always had...I just didn't get into detail about it because I didn't want them to worry. |
|  |  | Patient 26: You're not supposed to have stress because that's not healthy. |
|  |  | Patient 29: I got aches and pains like any other old man, but what are you going to do?<br>Interviewer: Those aren't things that you feel like you want to talk to your team about here?<br>Patient 29: No. Not here for sure because they're all extracurricular activities. |
|  | Centering patient values and self-advocacy | Patient 31: This is what husband's choice is. And we're not going to push that to say he should be on dialysis, but he chose not to. That should be the patient's choice, which the doctors totally respect. |
|  |  | Care Partner of Patient 110: He has a high potassium this visit, and they thought about adjusting his medicine. And we talked about just trying a diet for a while and then retesting the potassium. And I was totally fine with her to kind of circle around and do something completely different and see if it works. |
|  |  | Patient 143: I usually tell the physician whatever is bothering me or anything I'm curious about. I mean, my previous physician, I mean, he was very good. I mean, he answered any question I ever had. And any concerns like he knew I was paranoid about taking medication. So I always asked him a lot of questions about, "Can I take this with that?" And he let me message him at any time. |
|  |  | Patient 151: When I don't understand things, I tend to go like, "Okay. What's that?" And that's something I had to learn. But I'm sure clinicians have guided me through that beforehand without realizing I had to advocate for myself and kind of spoon fed me stuff. But I feel five years into it. I know more about asking and not smiling and nodding and walking away. |
| Theme 4: Physical symptoms and lifestyle changes were common but often attributed to comorbid conditions rather than CKD. | Fatigue and its impact on daily functioning | Patient 111: You used to grow a garden and we had a big garden. It has changed our life. As it's advanced, we've had to give up a lot of our normal activity. I used to do a lot of stuff. |
|  |  | Patient 8: Well, overall, I'm not as active as I used to be. I can't go out and cut trees down. And, you know, I can't raise cattle anymore. I had to get rid of them. |
|  |  | Patient 31: He gets tired you know easily, but I think that's one of the struggles that we started with in the beginning, is that he couldn't really do things like he used to be able to do and get them done in an hour. Now it takes him like four hours to do something that used to take him an hour. |
|  |  | Patient 110: I'm not capable of having a full-time job. I mean, I could try, but I couldn't be reliable. My energy levels up and down. Yeah, all over the place, for sure. Work my butt off one day and it takes two days to recoup. |
|  |  | Patient 143: My energy level, I really don't have an energy level. Like I could sleep 12 hours and still feel tired. I'm very tired all the time. I'm fatigued all the time. I don't know if it's chronic fatigue syndrome or what it is, but I always feel tired. And I have to make myself get up, like, out of bed or to like going to work is a struggle. Like, sometimes I work 16 to 12 hours a shift. |
|  |  | Patient 151: It's a quiet, boring existence, which I'm okay with. You know I don't have that many hobbies as I used to because of my lack of energy. |
|  |  | Patient 16: Once I come down sick, I kind of like lost all my friends I hung out with because I'm not as active as they are. |
|  | Dietary restrictions as an ongoing reminder of CKD | Patient 6: I don't think it has except for you know the sodium intake. I read every can. |
|  |  | Patient 31: Because it was more learning the new tools like what you could eat, what you couldn't eat, what you had to watch the sodium, watch the potassium. But now it's gotten to be like second nature. I used to be a big salt eater. No salt anymore. It took me a while to get used to it, I'll tell you. |
|  |  | Patient 143: I mean, like if it's in front of you, you'll probably want to eat it. Don't buy it. You won't eat it. You know I don't know. Yeah, like at my workplace, it's a lot harder because I work at an assisted facility where that's all they have is like, say, like fried foods. |
|  |  | Patient 151: Part of that initial coming into CKD is just like, "Okay, you need to tweak what you do, how you do it, what you take." I've been a soda hound since I was small. |

