## Appendix 1 for "Normalized barriers and unaddressed concerns: A qualitative study of the lived experiences of adults living in rural areas with advanced chronic kidney disease"

**Appendix 1:** COREQ (COnsolidated criteria for REporting Qualitative research) Checklist

| **Topic** | **Item No.** | **Guide Questions / Description** | **Reported on Page No.** |
| --- | --- | --- | --- |
| Domain 1: Research team and reﬂexivity | | |  |
| Personal characteristics | | |  |
| Interviewer/  facilitator | 1 | Which author/s conducted the interview or focus group? | Page 2 |
| Credentials | 2 | What were the researcher’s credentials? E.g. Ph.D., MD | Page 2 |
| Occupation | 3 | What was their occupation at the time of the study? | Page 2 |
| Gender | 4 | Was the researcher male or female? | Page 2 |
| Experience and training | 5 | What experience or training did the researcher have? | Page 2 |
| Relationship with participants | | |  |
| Relationship established | 6 | Was a relationship established prior to study commencement? | Page 2 |
| Participant knowledge of the interviewer | 7 | What did the participants know about the researcher? e.g. personal goals, reasons for doing the research | Page 2 |
| Interviewer characteristics | 8 | What characteristics were reported about the interviewer/facilitator? e.g. Bias, assumptions, reasons and interests in the research topic | Page 2 |
| Domain 2: Study design | | |  |
| Theoretical framework | | |  |
| Methodological orientation and Theory | 9 | What methodological orientation was stated to underpin the study? e.g. grounded theory, discourse analysis, ethnography, phenomenology, content analysis | Page 3 |
| Participant selection | | |  |
| Sampling | 10 | How were participants selected? e.g. purposive, convenience, consecutive, snowball | Page 2 |
| Method of approach | 11 | How were participants approached? e.g. face-to-face, telephone, mail, email | Page 2 |
| Sample size | 12 | How many participants were in the study? | Page 4 |
| Non-participation | 13 | How many people refused to participate or dropped out? Reasons? | Page 2 |
| Setting | | |  |
| Setting of data collection | 14 | Where was the data collected? e.g. home, clinic, workplace | Page 2 |
| Presence of non- participants | 15 | Was anyone else present besides the participants and researchers? | Page 2 |
| Description of sample | 16 | What are the important characteristics of the sample? e.g. demographic data, date | Page 4 |
| Data collection | | |  |
| Interview guide | 17 | Were questions, prompts, guides provided by the authors? Was it pilot tested? | Page 3 |
| Repeat interviews | 18 | Were repeat interviews carried out? If yes, how many? | Page 2-3 |
| Audio/visual recording | 19 | Did the research use audio or visual recording to collect the data? | Page 2 |
| Field notes | 20 | Were ﬁeld notes made during and/or after the interview or focus group? | Page 2 |
| Duration | 21 | What was the duration of the interviews or focus group? | Page 2 |
| Data saturation | 22 | Was data saturation discussed? | Page 3 |
| Transcripts returned | 23 | Were transcripts returned to participants for comment and/or correction? | Page 3 |
| Domain 3: analysis and ﬁndings | | |  |
| Data analysis | | |  |
| Number of data coders | 24 | How many data coders coded the data? | Page 3 |
| Description of the coding tree | 25 | Did authors provide a description of the coding tree? | Page 3 |
| Derivation of themes | 26 | Were themes identiﬁed in advance or derived from the data? | Page 3 |
| Software | 27 | What software, if applicable, was used to manage the data? | Page 3 |
| Participant checking | 28 | Did participants provide feedback on the ﬁndings? | Page 3 |
| Reporting | | |  |
| Quotations presented | 29 | Were participant quotations presented to illustrate the themes/ﬁndings? Was each quotation identiﬁed? e.g. participant number | Pages 5-9 |
| Data and ﬁndings consistent | 30 | Was there consistency between the data presented and the ﬁndings? | Page 3 |
| Clarity of major themes | 31 | Were major themes clearly presented in the ﬁndings? | Pages 5-9 |
| Clarity of minor themes | 32 | Is there a description of diverse cases or discussion of minor themes? | Pages 5-9 |

Developed from: Tong A, Sainsbury P, Craig J. Consolidated criteria for reporting qualitative research (COREQ): a 32-item checklist for interviews and focus groups. International Journal for Quality in Health Care. 2007. Volume 19, Number 6: pp. 349 – 35
