## Appendix 2 for "Normalized barriers and unaddressed concerns: A qualitative study of the lived experiences of adults living in rural areas with advanced chronic kidney disease"

**Appendix 2:** ADVANCED CHRONIC KIDNEY DISEASE (CKD) AGENDA SETTING PATIENT INTERVIEW GUIDE

Qualitative research is inherently iterative, so we may adapt this guide as our interviews progress.

Hi! I’m ______, thank you so much for taking the time to talk to me today.

We want to get your thoughts on your experience with Advanced Chronic Kidney Disease, also known as CKD. What you, and other people like you, say will help us create a tool to improve communication between people with advanced CKD and their doctors during outpatient appointments.

Questions: Do you have any questions for me before we begin?

Recording: We would like to record this conversation so that we can make sure we don’t miss anything you say. If you agree to this, we will remove anything that could identify you by name when we make the recording into a written document. Are you comfortable with me recording our conversation?

INTRODUCTION

Introduction: We’re working to make a tool called an agenda-setting intervention. The purpose of the tool is to help make communication between patients and their clinicians - people like doctors and nurses - easier. This will allow you to better advocate for yourself as a patient, share your needs with your doctor, and get the most out of each appointment. We want to make this tool specifically for patients with advanced CKD. That means chronic kidney disease stages 4 and 5.

Today, we want to hear more about your experience living with advanced CKD so that we can better understand how CKD impacts your life, and what kinds of things might be most helpful to you.

First, we’ll start off by asking you some questions about your overall experience living with advanced CKD.

Broad: Could you tell me a little about yourself (things like your hobbies, relationships, family, work…)?

- Is there anything else you would like to share about your healthcare journey?
- [probe if not mentioned:] When were you diagnosed with CKD?

Specific:

- How has your CKD affected your life (things like work, family, relationships, home, personal goals, financial wellbeing)?
- Could you describe a typical day in your life?
  - [probe if not mentioned:] Does CKD affect your day-to-day life? How so?
- Do you feel like you have a good understanding of your disease and condition?
  - How do you feel about the current management of your CKD?
- Do you have any worries about your CKD? If so, what?

RURALITY

Living in a rural area can make managing CKD more difficult; we’re curious about this for you.

- How do you get to your doctor's appointments and how long does it take to get there?
- How difficult is it for you to get to your doctor’s appointments?
  - [probe if not mentioned:] Does the travel time impact your ability to get the care you need?
  - [probe if not mentioned:] Have you missed appointments before? If so, why?
  - [probe if not mentioned:] How do you schedule your daily life around getting the care you need?
- Where do you get your medication from?
  - Do you encounter challenges?
  - [probe if not mentioned:] Does the travel time impact your ability to access your medication?
  - [probe if not mentioned:] What about medication delivery?
- Do you worry about being able to get your medication?
- [probe if not mentioned:] When you think about the challenges you just told me about, what would make getting your medication easier?

Appointments

Next, we want to talk about what your appointments with your doctors look like.

- Appointment structure:
  - Can you describe what it looks like from when you arrive through to the end of the appointment?
    - [probe if not mentioned:] How does your clinician start or set your appointments up?
    - What topics are typically discussed?
- During appointments:
  - What is the communication like between you and your doctor during appointments?
    - Do you feel like your clinician knows what matters to you?
    - Do you feel comfortable telling your clinicians what you want to discuss or expressing your opinions?
    - Do you feel heard when talking with your clinicians?
    - What about the other people on the care team? Like nurses, dieticians etc.
      - If any of these are answered with no, not comfortable etc. probe: Why? What makes you feel [uncomfortable etc.]? What might help you feel more [comfortable]?
- End/post-appointment:
  - How do you feel when you leave your appointments?
  - Do you feel like you cover everything you wanted to talk about by the end of your appointments?
  - When you leave an appointment, do you feel like you have a good understanding of your CKD?
    - What about your treatment plan?
  - [probe if not mentioned]: Do you feel there is a clear plan for next steps when you leave your appointments?

AGENDA SETTING

As we said, we are talking to you today so that we can better understand your experience with CKD. This is so that we can make a tool that will help improve communication between you and your doctor, so that patients like you can receive better care. This tool will help with something called agenda-setting. Agenda-setting is a process where a doctor works together with a patient to come up with a shared list of topics to talk about during the appointment. This could be as simple as your doctor starting the appointment by asking you, “what do you want to talk about today?”

- Follow-up with:
  - Does your doctor do agenda setting during your appointments? Could you tell me about what that looks like?

CKD TOPICS (CHRONIC KIDNEY DISEASE TOPICS)

So, as I mentioned, our team is designing an agenda setting tool. We want to design this tool specifically for the needs of patients with advanced CKD. We'd like to hear your thoughts on our tool so we can make sure it works for patients and care partners like you.

Please take a look at this agenda-setting tool developed for patients with chronic kidney disease. [ present CKD TOPICS now; use actual print outs]

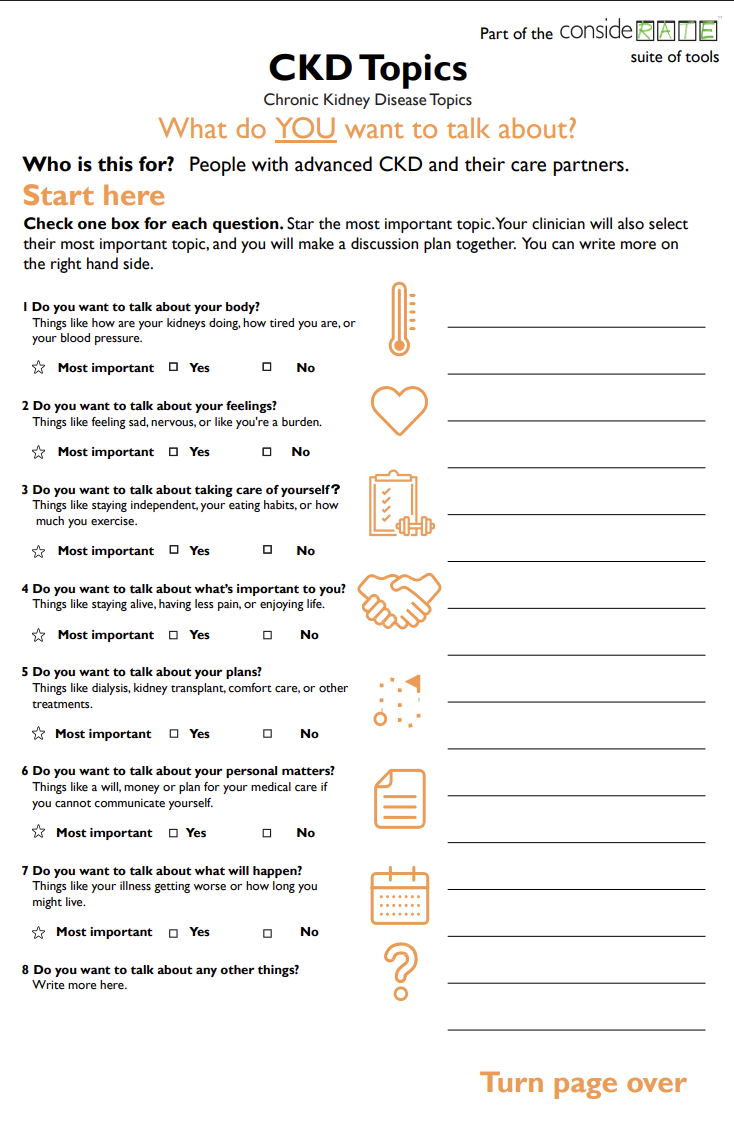

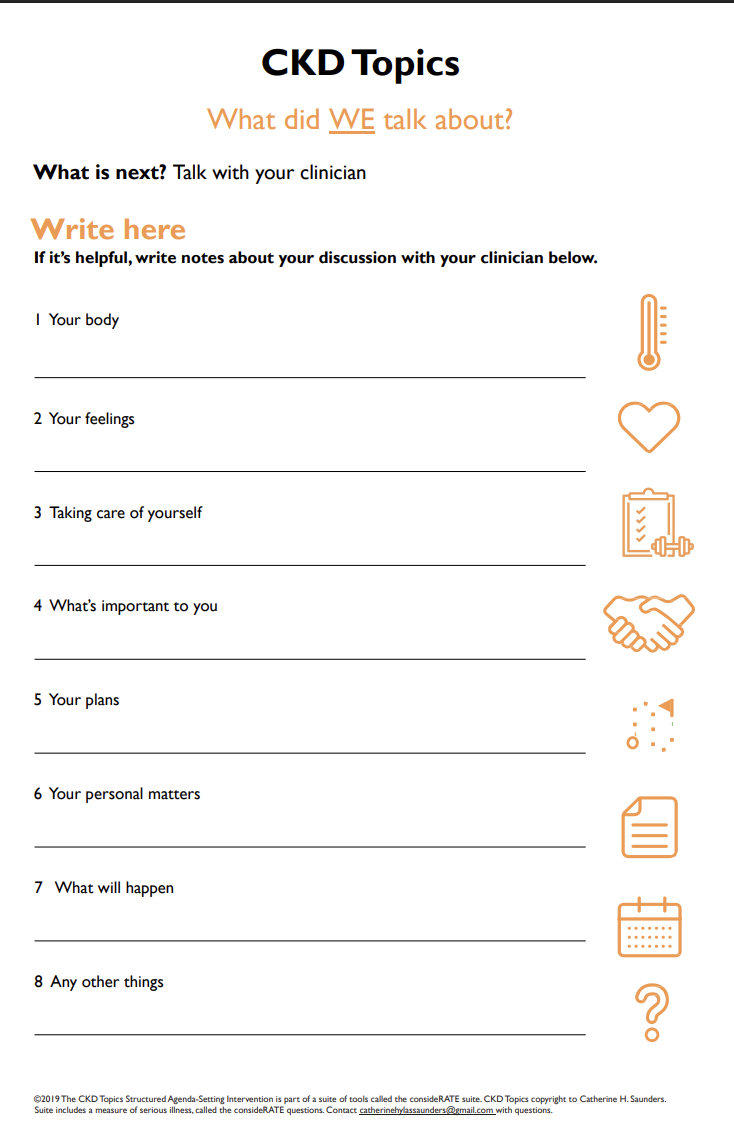

- What is your initial reaction to this tool?

Now we'll go through the tool line-by-line. I want to understand what you are thinking as you read.

Could you please read the instructions?

- What does that mean to you? In your own words?
- Are any words unclear?
- How could we make it clearer?
- Do you think the options: Yes, No, and star for most important make sense?
  - Do you like these options? Why or why not?
    - [Probe if necessary]: what would you change?

Could you please read question [x]. [repeat for every question]

- What does that mean to you? In your own words?
- Are any words unclear?
- How could we make it clearer?
- What about the examples, "things like"
  - What do they mean to you? In your own words?
  - Are we missing any examples you would include here
  - Are any words unclear?
  - How could we make it clearer?
- When you get to “What’s important to you?” ask specifically: do you think the examples should include “your spirituality” or “your family or relationships”?

Can you please read the instructions on the back page?

- What does that mean to you? In your own words?
- Are any words unclear?
- How could we make it clearer?
- What about the examples, "things like"
  - What do they mean to you? In your own words?
  - Are any words unclear?
  - How could we make it clearer?

- How helpful do you think this tool would be in appointments with your doctor?
  - [Probe if not included:] What do you like?
  - [probe if not included:] What do you not like?
- What about the way it looks?
- What about the topics listed on the tool?
  - [Probe if not included:] Do you think there are any missing?
  - [Probe if not included:] Do you think there are any that should be removed?
  - [Probe if not included:] Do you think there are any that should be changed?
- Do you have any concerns about using this tool? If so, what are they?
- Do you have any other ideas or feedback to share?

IMPLEMENTATION:

Along with your thoughts on the tool itself, we would also like to know your opinion on how we can best use an agenda-setting tool.

How do you think an agenda-setting tool like this could fit into your care?

- Imagine you're using the agenda-setting tool. When do you think you would want to fill it out?
  - Before the appointment at home or before you get to the hospital?
  - In the waiting room?
  - During your conversation with your clinicians?
- How do you think this tool should be delivered to patients?
  - Online/ Website/MyDH
  - Printed
  - Sent through app/email
  - Discuss with clinicians
- How frequently do you think you would fill out this tool?
  - Before every appointment?
  - At certain times in the treatment course?
    - If so, which times?
  - Other times?
- Do you want the option to revisit or discuss this tool during each visit?
  - If not every visit, how often?
- How much time should it take to use an agenda-setting tool?
  - How much time do you think should be for filling out the tool?
  - How much time do you think should be spent talking with your clinicians about the tool?
- Is there anything we haven’t talked about that you think I should know?

Thank you so much for taking the time to speak with us. We really value your perspectives and input in designing a tool that hopefully works for patients with CKD and their care partners. Here is a thank you letter.

If this is something you’d like to stay involved in, we’d love to hear your opinion and there may be future research involvement opportunities for patient partners. You can reach out to the folks on the thank you letter!
