## Appendix 3 for "Normalized barriers and unaddressed concerns: A qualitative study of the lived experiences of adults living in rural areas with advanced chronic kidney disease"

**Appendix 3:** PATIENT DEMOGRAPHICS

Questions about you

Please answer the following questions about you, your experiences and your family. Feel free to skip any questions that you would prefer not to answer.

1. **What is your age?**

[REDCAP drop-down options]

2. **What is your gender?** Please choose one.

- Female
- Male
- Non-binary
- Other

3. **Which groups do you most closely identify with?**

Please choose all that apply.

- American Indian or Alaska Native
- Asian
- Black or African American
- Native Hawaiian or Other Pacific Islander
- White or Caucasian
- Other (please specify): _____________

4. **Are you Spanish, Hispanic, or Latino/a?** Please choose one.

- Yes
- No

5. **Why are you visiting the clinic today?** Please choose all that apply.

- Chronic Kidney Disease (CKD)
- Kidney transplant or follow-up
- Other: ____________

6. **Do you know your glomerular filtration rate (GFR)? This is sometimes used to measure kidney health.** Please choose one.

- Stage 1 with normal or high GFR (GFR > 90 mL/min)
- Stage 2 Mild CKD (GFR = 60-89 mL/min)
- Stage 3A Moderate CKD (GFR = 45-59 mL/min)
- Stage 3B Moderate CKD (GFR = 30-44 mL/min)
- Stage 4 Severe CKD (GFR = 15-29 mL/min)
- Stage 5 End Stage CKD (GFR < 15 mL/min)
- Unsure

7. **Do you have kidney disease?** Please choose one.

- Yes
- No
- Unsure

8. **(skip logic) How long have you known you had kidney disease?**

[REDCAP drop-down options]

9. **Are you on dialysis?** Please choose one.

- Yes
- No
- Unsure

10. (skip logic) **What kind of dialysis are you on?** Please choose all that apply.

- In-center hemodialysis (in healthcare facility)
- Home hemodialysis (at home)
- Peritoneal dialysis (through belly button)
- Unsure

11. **Do you have any of these health conditions?**

Please choose all that apply.

- Hypertension
- Cardiovascular disease
- Diabetes
- Other, please specify: ____________

12. **What is your primary language?** Please choose one.

- English
- Spanish
- French
- Mandarin Chinese
- Other, please specify: _____________

13. **Are you religious or spiritual?** Please choose one.

- Very religious/spiritual
- Moderately religious/spiritual
- A little religious/spiritual
- Not at all religious/spiritual

14. **How would you describe the place that you live?** Please choose one.

- Urban (in a large city)
- Suburban (near a large city)
- Rural (not near a large city)
- Very rural (very far from a large city)

15. **How long do you have to travel to see the primary clinician that helps you manage your kidney disease?** Please choose one.

- 15 minutes or less
- 16 to 30 minutes
- 31 to 60 minutes
- Longer than 60 minutes

16. **What zip code do you live in?**

[REDCAP Drop down]

- _______________

17. **How confident are you in filling out medical forms by yourself?** Please choose one.

- Extremely
- Quite a bit
- Somewhat
- A little bit
- Not at all

18. **What is your highest level of education?** Please choose one.

- Never attended high school
- Some high school, no diploma received
- High school diploma or equivalent (e.g. GED)
- Some college, no degree received
- Associate’s degree)
- Bachelor’s degree
- Master’s degree
- Doctoral degree

19. **To the best of your knowledge, what is the current total yearly income in your household? This includes all salaries, wages, profits and other income before taxes or deductions for anyone 15 or older that lives with you.** Please choose one.

- Less than $25,000
- $25,000 to $34,999
- $35,000 to $49,999
- $50,000 to $74,999
- $75,000 to $99,999
- $100,000 to $149,999
- $150,000 to $199,999
- $200,000 or more

20. **What is your primary source of income?** Please choose one.

- Social security
- Full-time employment
- Part-time employment
- A household member’s full-time employment
- A household member’s part-time employment
- Savings
- Other, please specify: ____________

21. **How many dependents are living or staying in your household? This includes you, your legal spouse, and any tax dependents. For example, if you live with your spouse and have a child who is dependent on you, you would select 3.** Please choose one.

- 1
- 2
- 3
- 4
- 5
- 6
- 7
- 8
- 9
- 10
- 11
- 12+

22. **Do you have health insurance?** Please choose one.

- Yes
- No

23. (skip logic) **What kind of health insurance do you have?**

Please choose all that apply.

- Provided by employer
- Purchased yourself
- Medicare
- Supplemental
- Medicaid
- Other
